# A Prospective Study of Outcomes After Tubularized Incised Plate (TIP) Urethroplasty: a Multivariable Analysis of Prognostic Factors in Children

**DOI:** 10.1101/2020.09.15.20193037

**Authors:** M. Reza Roshandel, Fahimeh Kazemi Rashed, Tannaz Aghaei Badr, Samantha Salomon, Mohammad Seyyed Ghahestani, Fernando A. Ferrer

**Affiliations:** Department of Urology, Icahn School of Medicine at Mount Sinai, New York, NY, USA, Correspondence (present address): 1425 Madison Avenue New York, NY; 10029; Tabriz University of Medical Sciences, Iran; Clinilabs Drug Development Corporation, New York, NY; Tehran University of Medical Sciences

**Keywords:** Hypospadias, TIP, Urethroplasty, outcomes, Prognosis, Prospective study, Chordee

## Abstract

**Background:** Tubularized incised plate (TIP) urethroplasty as the most common hypospadias repair method, aims to achieve normal functioning of the penis along with cosmetic reconstruction. However, there are remaining questions toward anatomical prognostic factors affecting the results of surgery. Lack of age-matched controls or controlling for meatal location, employment of several surgical techniques or multiple surgeons, or age heterogeneity of the study population are the problems affected the results of the current body of literature.

**Objective:** This prospective study aimed to evaluate the preoperative factors to predict future complications associated with hypospadias repair outcomes in males aged between 1-3 years and performed by a single surgeon with employing multivariable analysis.

**Patients and methods:** A prospective cohort of 101 males aging from 1 to 3 years with distal to mid-shaft hypospadias were consecutively selected for TIP repair. The urethral plate dimensions in erect and flaccid states, penile length, glans diameter, and chordee were evaluated individually before reconstruction. After surgery and during follow-up visits, the subsequent transient and persistent complications were recorded.

**Results:** Postoperatively, the acute transient events were observed in 42 cases (41.6%) and the persistent complications in 16 cases (15.8%). The uncomplicated group had a higher percentage of patients with distal meatal location than the complicated group (P=0.01%). Furthermore, fistula formation was notably higher in the group with acute surgical site infection (P<0.001). wedThe analysis also shothe width of the urethral plate to be associated with the development of complications (P=0.03).

**Conclusion:** By performing TIP by a single surgeon on a homogenous study population and eliminating the impact of severe chordee as a potential cofounding variable, this study prospectively found that out of the anatomical specifications, pre- and postoperative factors, the urethral meatus location was the only significant and independent predictor of the development of complications in young children with midshaft to distal hypospadias. Moreover we found that in young children the wider the plate was, the more complications happened. Consequently, we hypostatized that in young children who their anatomical dimensions are almost in same range of values, a combination of urethral width and depth should be considered in the investigation of prognostic factors for hypospadias repair outcomes. (figure 1)

## INTRODUCTION

As a common congenital anomaly, hypospadias typically requires surgical repair during the early years of life. Hypospadias is a consequence of defective closure of the urethral fold and is characterized by deficient ventral preputial skin, a more proximally located meatus which can be accompanied by the chordee. Despite, the fact that numerous procedures have been described, today, the most common repair method for distal hypospadias is the TIP (Tubularized Incised Plate) urethroplasty [1].

**Figure 1.**
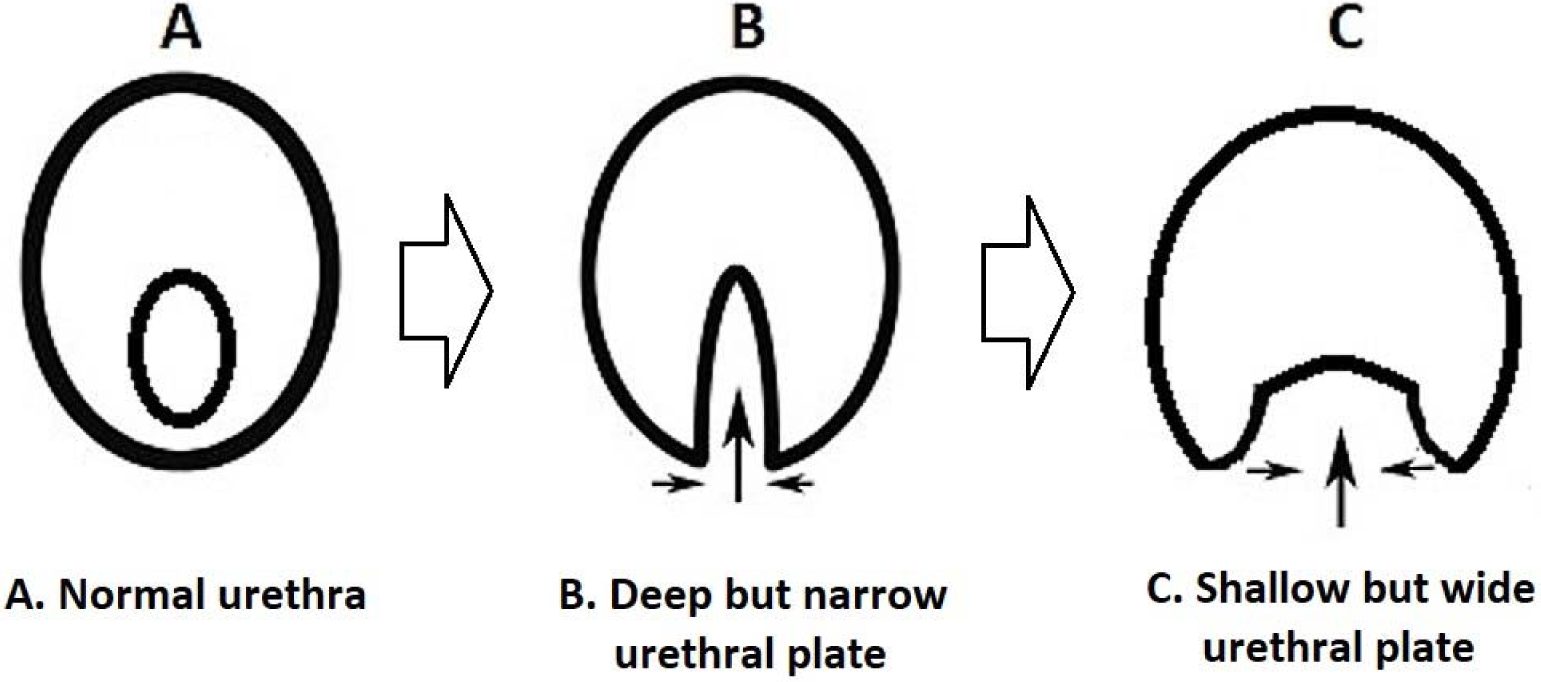
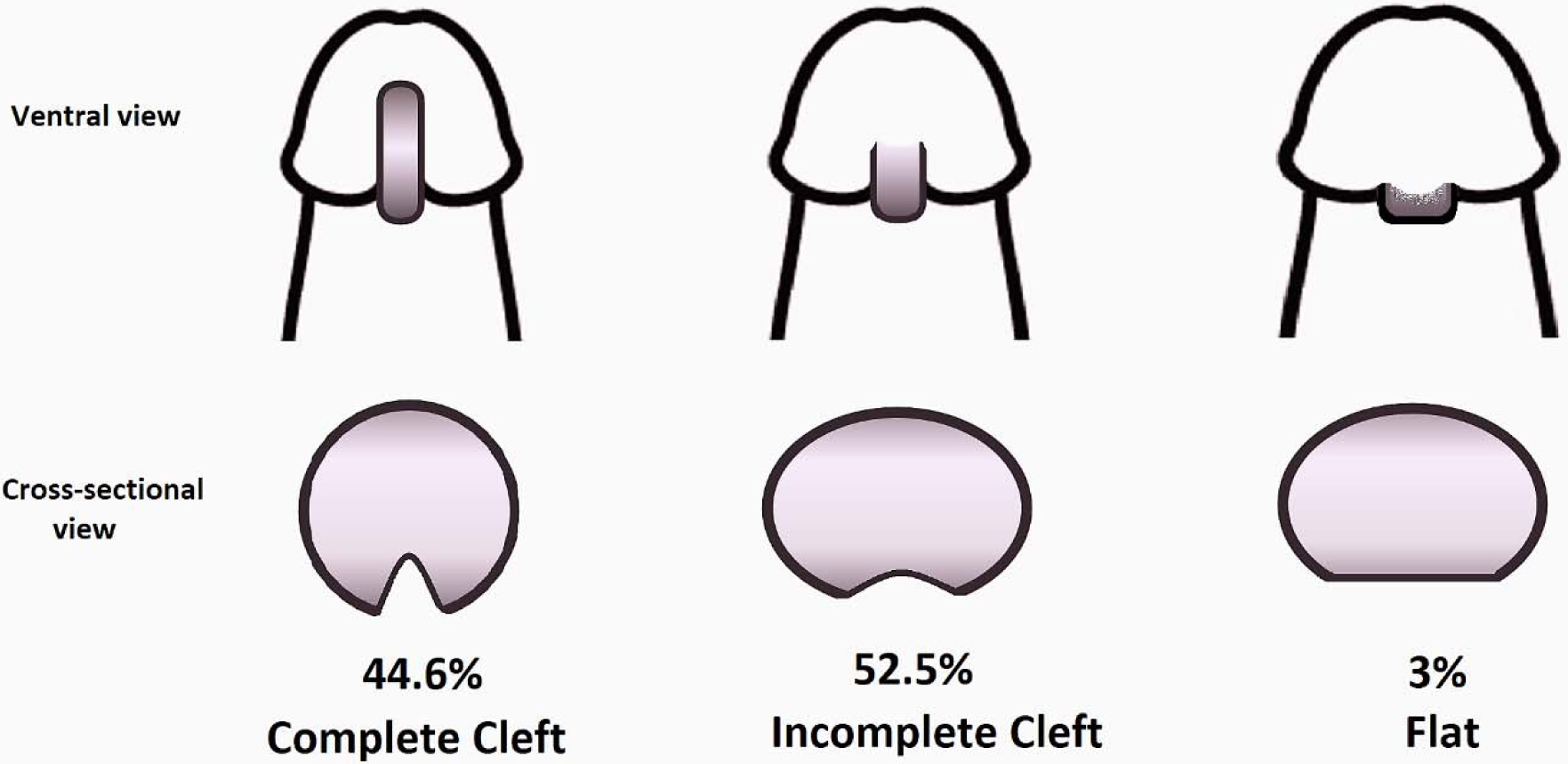
Glans types according to the glanular urethral plate cleft and the prevalence in the study.

Fistula, meatal/urethral stenosis, and dehiscence are amongst the most common complications following primary repair of hypospadias [2, 3]. A central question has been the preoperative or intraoperative factors that can predict future complications. Identifying these parameters would allow risk assessment and potentially lead to new techniques to circumvent these adverse features.

Wide discrepancy exists in the literature regarding prognostic factors related to the urethral plate and other penile dimensions. For example glans and urethral plate width have been correlated with postsurgical complications. However useful prognostic factors in the management of hypospadias remain poorly defined [1, 3-8]. Factors complicating prior studies include, non-homogenous patient populations, variations between multiple surgeons, lengthy accrual times during which subtle technique changes may have occurred. In this study, we assess several possible prognostic factors for complications in a rigorous, homogenous series of de-novo hypospadias cases, 1-3 years in age.

## MATERIALS AND METHODS

In this study, 101 children with hypospadias were evaluated in a prospective cohort study concerning the characteristics of their urethral plate dimensions in erect and flaccid states, penile stretched length, glans characteristics, and chordee. Subsequent, acute events and postoperative complications during the follow-up period were carefully monitored and recorded by collecting the data with the help of a survey form (Supplemental file).

The study received the institutional review board approval (IRB No. 92/3-1/4) and was conducted at the urology clinic of a referral hospital within 12 months.

The inclusion criteria were the first time surgery cases of distal to mid-shaft hypospadias with an age between 1 to 3 years old.

The exclusion criteria were the co-existence of moderate to severe chordee, micro-penis, or a prior history of hypospadias surgery or circumcision. Micropenis was defined as a dorsal length of a stretched penis less than 1.9 cm in length for a term infant or if the length is less than 2.5 standard deviation below the mean. Chordee or ventral curvature (VC) was recorded at the first visit and also after performing the erection test before making skin incisions. Chordee is defined as a penile shaft curvature and is categorized based on its degree: mild (<30), moderate (30–45), and severe (>45) curvature [9].

### Procedure

After the induction of anesthesia in the operation room, the diameter of glans, the length and width of the urethral plate, and the presence of ventral curvature were determined in both flaccid and erect states. Chordee was corrected by degloving and extensive ventral dissection if needed. All patients underwent TIP repair. The urethral plate was tubularized over 8 Fr Nelaton stent with a three-layer reconstruction. A ventral dartos flap was mobilized for additional coverage and secured on each side as the flap the 3^rd^ layer using 5-0 Vicryle interrupted stitches. The skin was closed by subcuticular continuous 6-0 Vicryl stitches. Two-layer closure of the glans wings was carried out for glanuloplasty. Additional coverage of the glandular urethra was provided by using a separate dartos flap for glanular urethral protection. We didn’t insist on the construction of a total glanular conical shape, to prevent the stricture of neomeatus, rather, we built the neomeatus approximately 3 millimeters below the normal location. As a final step, we replace 8 Fr stent with similar 6 Fr one. After the surgery, the patients were dressed, using Vaseline gauze and ordinary sterilized gauze. For about 2 to 4 days, they received IV cefazolin. The stent removed on between 5^th^ to 7^th^ day postoperatively. All surgeries were done by a single high volume attending pediatric urologist. In no cases was preoperative testosterone used. In the postoperative phase, emerging complications were divided into categories: transient acute events and persistent complications. Patients were revisited after 1 and 6 months. At the second follow-up visit at least one of the parents assessed satisfaction completing a survey list in terms of cosmetic outcomes (based on parents overall perception regarding meatal location and shape, and general cosmetic appearance) and functional outcomes (voiding without splitting, spraying or straining and away from the feet) with yes or no questions comparing their perception before and after repair.

## STATISTICAL ANALYSIS

The Data obtained is presented as standard deviation (±mean) and average (%). Data analysis was performed in SPSS version 24.0. The T-test was employed for independent groups to compare quantitative variables.

Ki2 or Fisher test was applied to compare qualitative data. Furthermore, A regression logistic model was deployed for multivariable data analysis and determining independent parameters. P values less than 0.05 were considered statistically significant.

### Results

One hundred and one patients were included. The mean age of the patients undergoing surgery was 19.86 ±8.60 (12-36) months. The location of urethral meatus was distal in 88 patients (87.1%) and mid-shaft in 13 patients (12.9%) (diagram 1). The mean dorsal length of the stretched penis was 22.99±4.03 millimeters (mm) (range: 15-32 mm). The median length of the penis at the ventral surface was 19.04 ±3.81 mm (range: 10-29 mm). Thirty-one patients had minor ventral curvature (30.7%). The glans urethral plate was categorized according to the depth of the urethral groove in the flaccid state and included a glans group with complete cleft with 45 (44.6%) cases, incomplete cleft, 53 cases (52.5%), and a flat group with 3 cases (3%) (Figure 1).

The mean width of the urethral plate was 9.51± 2.50 (4-20) mm in flaccid condition and 10.59±2.62 (5-21) mm in the erect state. Accordingly, the width of the urethral plate was less than 1 cm in 46 cases (45.5%) and 1 cm or more in 55 cases (54.5%). The mean length of UP was 10.52±2.88 (6-18) mm when erect and 11.86±2.41 (8-22) mm when flaccid. The mean diameter of glans was 17.18±3.08 (12-23) mm (when flaccid).

## POST-OPERATIVE FINDINGS

The mean length of the reconstructed urethral plate was 12.42±2.51 (8.20) mm. Acute postoperative events were observed in 42 cases (41.6%). The mean time to the development of acute events was 2.67±1.16 (1-5) days. Acute events (complications) are different from persistent complications in that they are transient. The mean total follow-up time was 6.58±3.36 (1-12) months. 85 (84.2%) patients did not have any long term complications. In contrast, 16 cases (15.8%) developed persistent complications (present please), the most prevalent of which was fistula observed in 5 cases (3%). Other complications included recurrent chordee, meatal stenosis, glans, urethral dehiscence, and ventral scarring of the glans (diagram 2 and Table 1). Severe immediate post-op edema was seen in 8 cases (7.9%) on the first after surgery. This could be because of manipulations in lymphatic drainage during dissections. Dysuria and UTI were reported in 23 (22.8%) and 2 cases (2%), respectively. Based on the research survey, surgical results were satisfying to 100 (99%) parents in terms of cosmetic and functional results. The study variables in the complicated and non-complicated groups are compared and summarized in Table 1. Width of the UP in the flaccid penis, length of UP in the erect penis, and location of meatus emerged as significant predictors of complications. However, UP width, in our study, was associated with poor outcomes. In all other measures, no statistical difference was observed (Table 1). Counterintuitively, the complicated group had a higher percentage of patients with the urethral plate width of ≥ 1 cm. (87.5% vs 48.2%, PV=0.004)

**Table 1.**
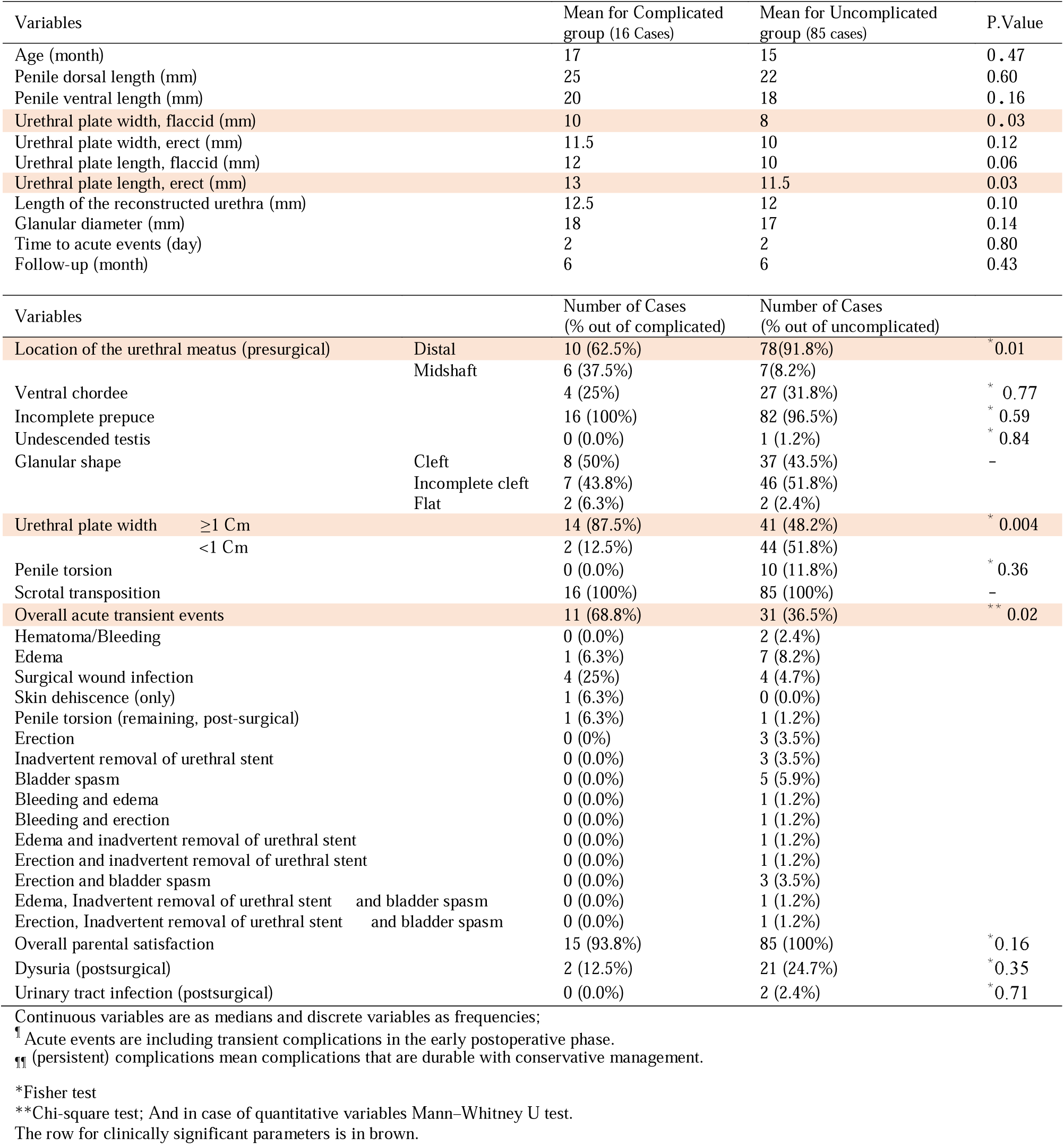
Pre-surgical findings and acute post-surgical events^¶^ in the complicated^¶¶^ (Group 1) and uncomplicated (Group2) patients

In the case of the qualitative variables, the uncomplicated group had a higher percentage of patients with a distal original meatal location than the complicated group (91.8% vs 62.5% respectively, P.V = 0.01%)

On multivariable analysis to find independent risk factors for complications, the width of the urethral plate was not significant (RR=0.94, P=0.61) when it was flaccid; neither was the length of the urethral plate (RR=0.9, P=0.93) when it was erect. Only the location of the urethral meatus (orifice) was significantly related to postoperative complications(RR=7.69, P=0.01). Post-operative acute events were not significantly associated with complications (RR=0.25, P=0.06). In patients with acute surgical site infection, 4 cases developed fistulas (50%), while in patients without acute infection, the fistula was observed in 2 cases (2.1%). Thus fistula formation was notably higher in the group with acute surgical site infection (P<0.001) (Diagram 3).

## DISCUSSION

Approximately 1 out of every 10 distal hypospadias surgeries develops a complication. Surgeons attempt to identify risk factors for complications to make modifications in techniques to improve their results [10]. In this study, we evaluated the results of the mid-shaft and distal hypospadias surgery in terms of potential factors predictive of complication. During 170 days of average follow-up time, late complications were noticed in 15.8% of the patients (Diagram 4). Studies conducted by Snodgrass et al [11], Aslam et al [12], Mane et al [13],Xu et al [14], and Springer [15], estimate the complications after TIP operation to be between 7 to 31%. o (Table 2). Thus, overall complications in our study (16%,) were consistent with other contemporary series.

**Table 2.** Complication rate in different studies

|  | Complication rate | Number of patients | Age range (mean) | Type of Hypospadias | Follow-up length (mean) |
| --- | --- | --- | --- | --- | --- |
| **Snodgrass et al** | 30% | 118 | 3-156 months | Proximal | 8 weeks - 82 months |
| **Aslam et al** | 7% | 93 | 3-7 years<br>(3.5 yr) | Midshaft and distal | 3 - 103 months<br>(56 m) |
| **Shivaji et al** | 11% | 100 | 1-5 years<br>(2.7 yr) | Distal | 3 months - 4 years<br>(23 m) |
| **Xu et al** | 20% | 176 | 1-14 years<br>(4.3 yr) | Proximal | 12-51 months |
| **Mane et al** | 11% | 100 | 1-5 years<br>(2.7 yr) | Distal | (23 m) |
| **Dokter et al** | 31% | 205 | 4.5 months–11.1 years<br>(10.3 months) | All | 1 month -13.5 years (4.7 years) |
| **Arlen et al** | 14.1% | 262 | 12.3± 3.7 months | All | 7.7±9.3 months |
| **Aboutaleb H.** |  | 100 | 9 months and 12 years<br>(4.3 years) | Distal | 2-24 months (6.4 months) |

### Meatal location

It has long been thought that the hypospadias with a distal meatal orifice, is less prone to complications than those with proximal defects [16-19].

Spinoit et al studied the results of hypospadias surgery in 474 patients. In the univariate logistic regression analysis, among the statistically significant predictors related to re-operation were hypospadias being proximal [20]. Conversely, Arlen et al showed the meatal location to have no significance on multivariable analysis. They concluded it’d be more important considering the entire hypospadias complex anatomy in defining severity, rather than the meatal location merely [21].

In our study, although on the single-variable analysis the length and width of the urethral plate, location of the meatal orifice, and acute events were significantly related to the complications. In the multivariable study, it was only the initial meatal location that was found to be statistically significant (p= 0.03) (Diagram 5). Accordingly, we concluded that the closer meatal orifice was to the coronal margin (as the threshold), the fewer complications existed (RR= 7.69, P=0.01). While the difference between complicated and uncomplicated groups regarding the meatal location was statistically significant (P=0.03), it is noted that the actual measured differences were small (1.5 in millimeters). This marginal difference wouldn’t be practically important from the technical point of view and it is difficult to envision how daily surgical practices would be modified.

### Width

Several studies have shown different results regarding the impact of the urethral plate width. In Holland and Smith’s study of 48 cases ages 3–7 years old, the width of the urethral plate was found to be directly associated with the development of a fistula. Their cut off point of urethral plate width for complications was found to be 8 millimeters [5]. Further studies by Sarhan et al, K.E. Chukwubuike et al, and Aboutaleb H confirmed this finding [22, 23]. In a study by Seleim significant connection between urethral plate width and complications was found with a cutoff value of less than 4 mm [24]. Arlen et al employed a scoring system called glans-urethral-meatus shaft (GMS) to evaluate the probability of hypospadias postsurgical events. This system was based on the size of glans, the specifications of the urethral plate, the meatal location, and the severity of shaft curvature. The proposed scoring system was able to anticipate the occurrence of post-surgery complications in this group of patients. Only the degree of chordee arose as an independent predictive factor in the multivariable study analysis. A major limitation of this study was its different reconstructive techniques were used [21]. Other studies have failed to find any significant connection between UP width and complications [25-27].

In summary, while previous studies have reported varying results about UP width, in our study the width was associated with the poor surgical outcomes in the single-variate analysis (RR=7.69, 0.004). However, it wasn’t significant in the multivariable study (P=0.61). It should be also noted that the width and depth of the urethral plate/groove are parameters that are connected, meaning that in a newborn with a urethral anomaly, the wider the ventral urethral groove is, the less depth it will have. So one may consider the width and depth of the urethral plate together (fig. 2). This could be an interesting topic for future investigations.

**Figure 2.**
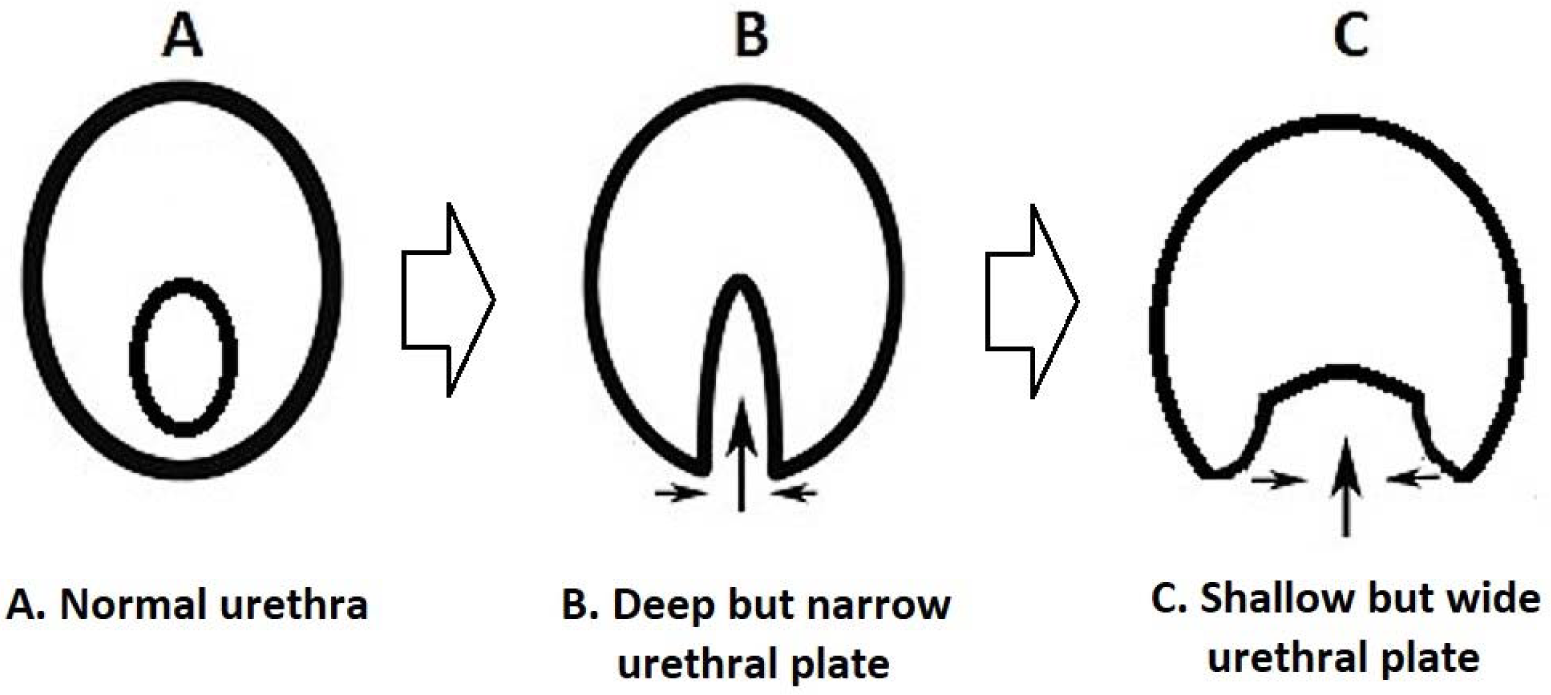
The inverse relationship between width and depth in hypospadias anomalies.

### Fistula and surgical site infection

Fistula developed in 6 cases and wound infection in 8 cases (Diagram 3). We found a significant relationship between fistula formation and surgical site infection (P<0.001). None of the fistula cases had meatal or urethral stenosis. Infection along with relatively low vascularity may result in poor wound healing and resultant fistula. So, finding measures to reduce the rates of infection as a modifiable risk factor could be considerably helpful.

### Glans

Although smaller glans widths are thought to be associated with greater difficulty and perhaps complications, our study along with some other studies, hasn’t shown glans diameter to be a risk factor for complications after TIP [26-29]. Also we found no significant differences in glanular groove shapes (complete cleft, incomplete cleft, flat) between two groups. This result correlates well with a study performed by da Silva et al [26]. Other studies focusing on distal hypospadias have found a significant difference related to glans size [30, 31]. It’s not clear if these different results are because of variations and modifications in glansplasty technique by surgeons [10].

### Chordee

In terms of superficial chordee, there was no significant difference between complicated and uncomplicated groups. Severe chordee cases were excluded from our study. Severe chordee (or penile curvature) can be indicative of underlying severe hypoplasia of corpus spongiosum, Dartos, or Buck’s fascia in the urethral area, which suggests the severity of hypospadias defect is more profound and blood supply may be more limited. For this reason, the existence of Severe chordee per se may be a major risk factor for complications particularly fistulas, and can affect other variables in the study [32]. Including Severe chordee (or proximal hypospadias) into variables may lead to undervaluing or obscuring the role of other potential risk factors. It seems that this would be a point missed in some studies and might be one of their main limitations. Our study primarily aimed to eliminate this confounding error.

It should be pointed out that in our study, the variables were measured and reported by a single observer. It has been shown that there are remarkable differences between observers in measuring the dimensions of the glans [21]. It is not crystal clear whether or not this limitation has been able to affect the results of the present study. The inter-observer variability measurements in future studies in this field will help.

Because our study group consisted of children 1-3 years old, the results might not be the same for other age groups. We recommend determining prognostic actors for different age groups separately.

## Conclusion

Although studies have been conducted by many authors in the hypospadias field, the current body of literature has been plagued by lack of age-matched patients, lack of controlling for meatal location, diversity in surgical techniques, surgeons’ multiplicity, or non-uniform distributions of study population ages. So, we tried to overcome these vulnerabilities by employing one repair technique, performed by a single surgeon, in all mid-to-distal hypospadias cases, with no substantial chordee, and enrolling a homogeneous study population. This prospective study, using multivariable analysis suggests that meatus location is the only significant predictor of the development of complications. However, this in an unmodifiable factor before surgery. We also found that surgical site infection may have an association with fistula formation later on.

We recognize that the follow-up period of 1 year could be considered short for hypospadias. Additionally, there is the inevitability of subtle anatomic variation in case series of this nature. Moreover, Last but not least, we propose considering a combination model of urethral depth and width with age-related adjustments for future studies as well as the inter-observer variability measurements. while we have focused on rigorously on standardizing multiple variables to ensure a homogenous population.

Moreover, we found that in young children the wider the plate was, the more complications happened. Consequently, we hypostatized that in young children who their anatomical dimensions are almost in the same range, a combination of urethral width and depth should be considered in the investigation of prognostic factors for hypospadias repair outcomes. Since measurement of depth of urethral plate is challenging even in older children, urethral plate width might be the single most important dimension of urethral representing the clinical situation.

## Supporting information

Mesurement . suplementary 1

Mesurement . suplementary 2

Mesurement . suplementary 1

## Data Availability

All data are availabe including the analyses.

## Abbreviations and Acronyms

TIP: Tubularized Incised Plate
VC: ventral curvature
SSI: surgical site infection

---

**Diagram 1.**
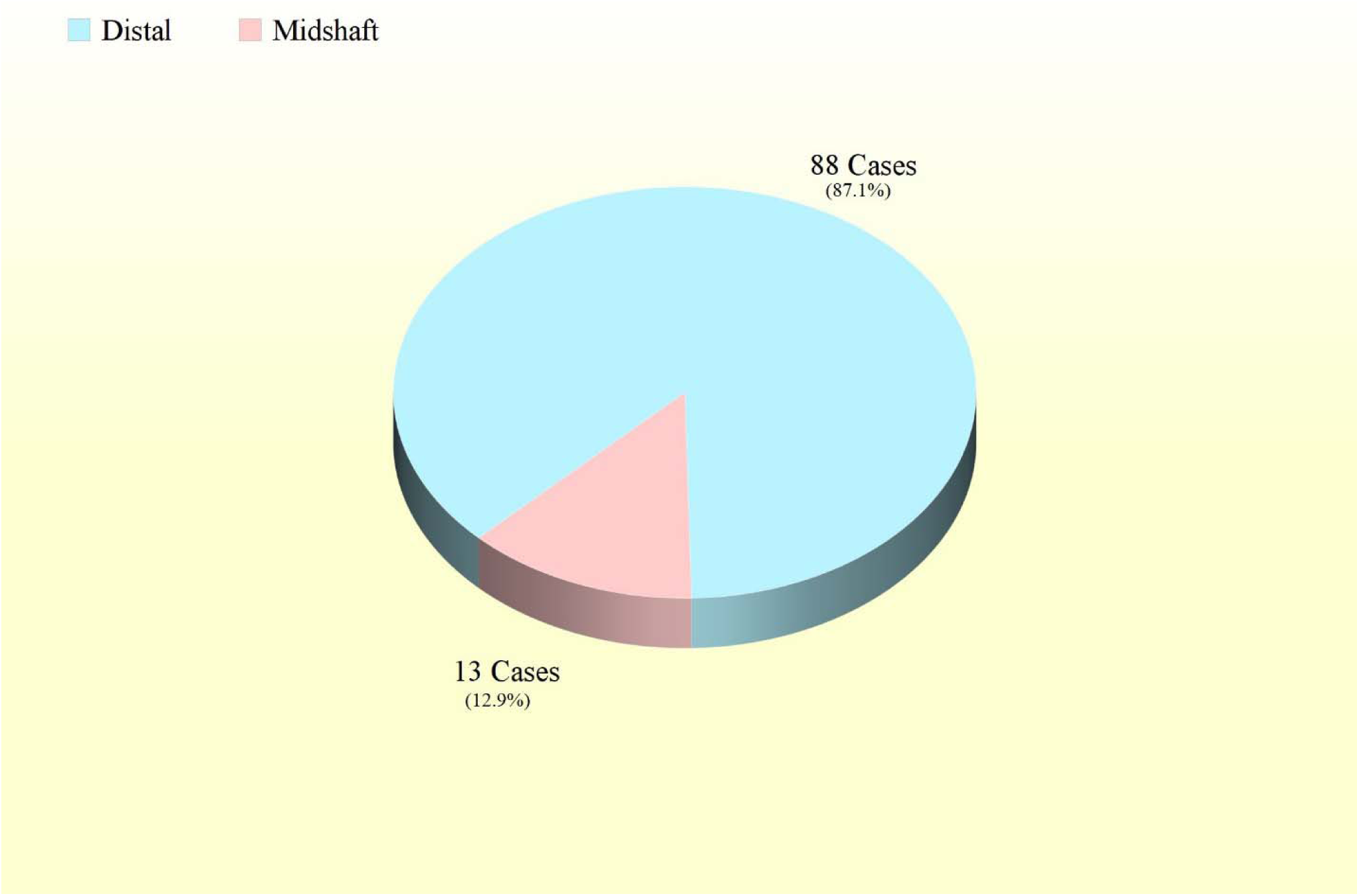
Meatal location: Prevalence and percent of study patients.

**Diagram 2.**
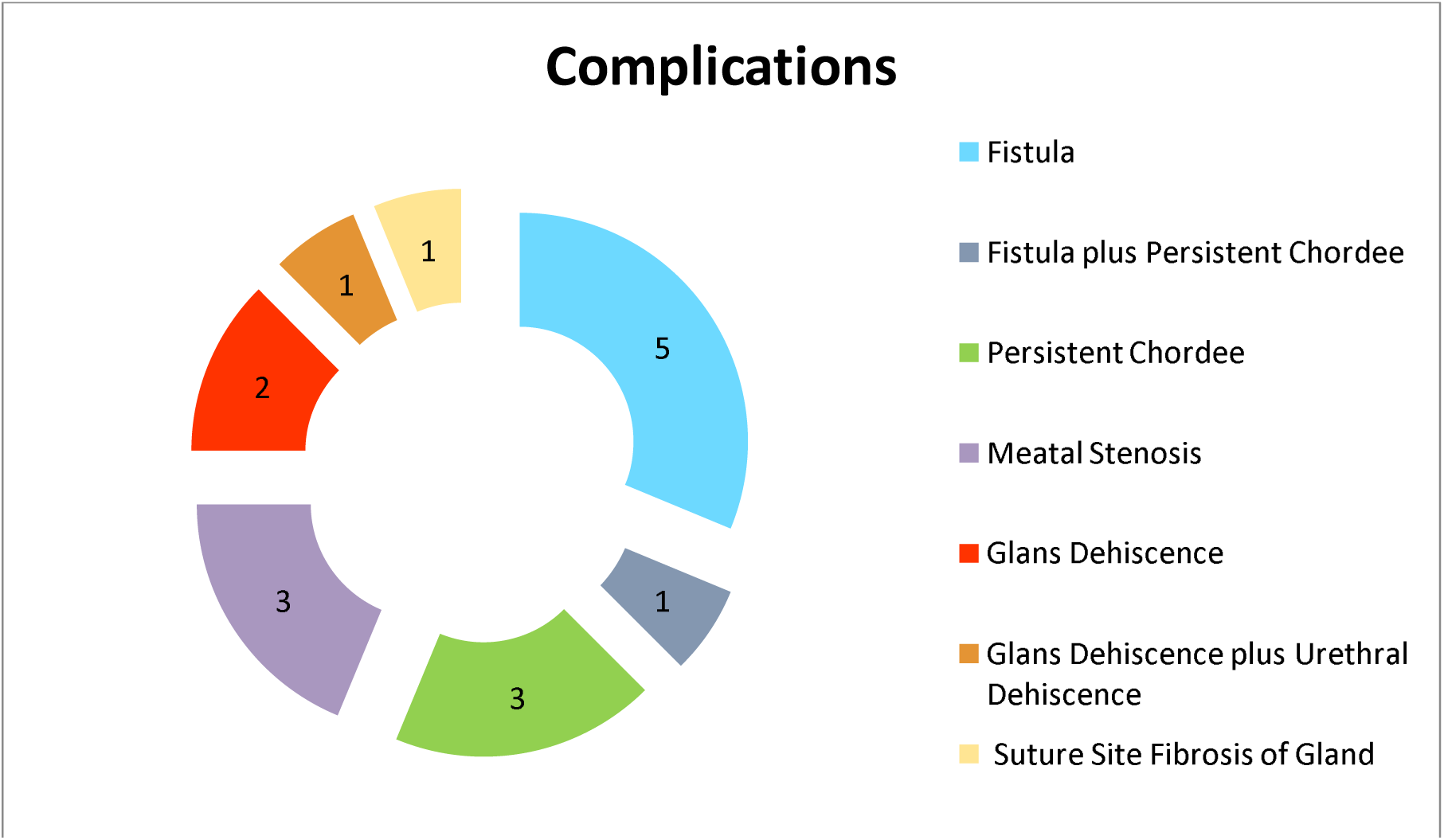
Doughnut chart of of complications and pertaining number of patients.

**Diagram 3.**
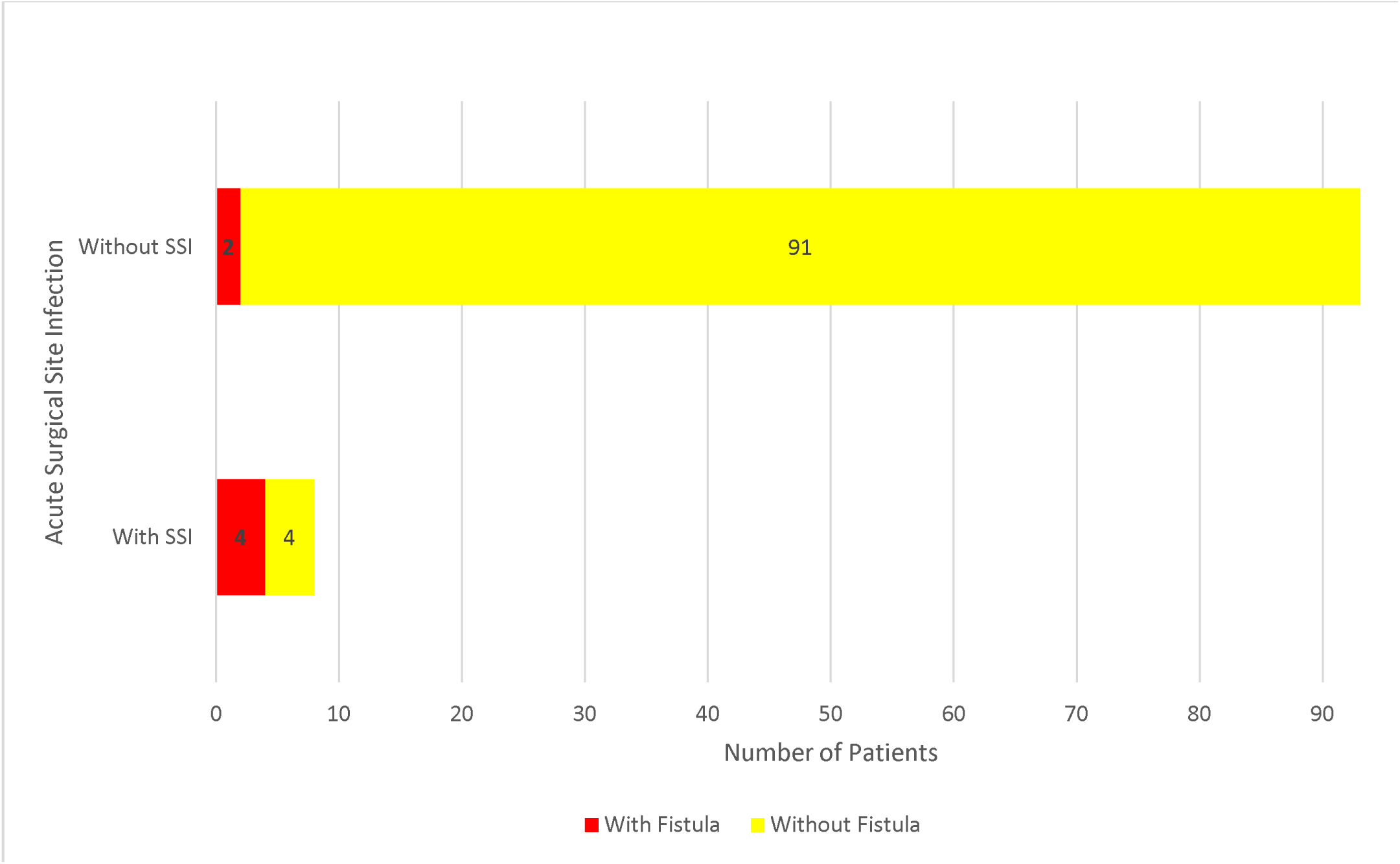
Surgical Site Infection and Future Fistula Developement.

**Diagram 4.**
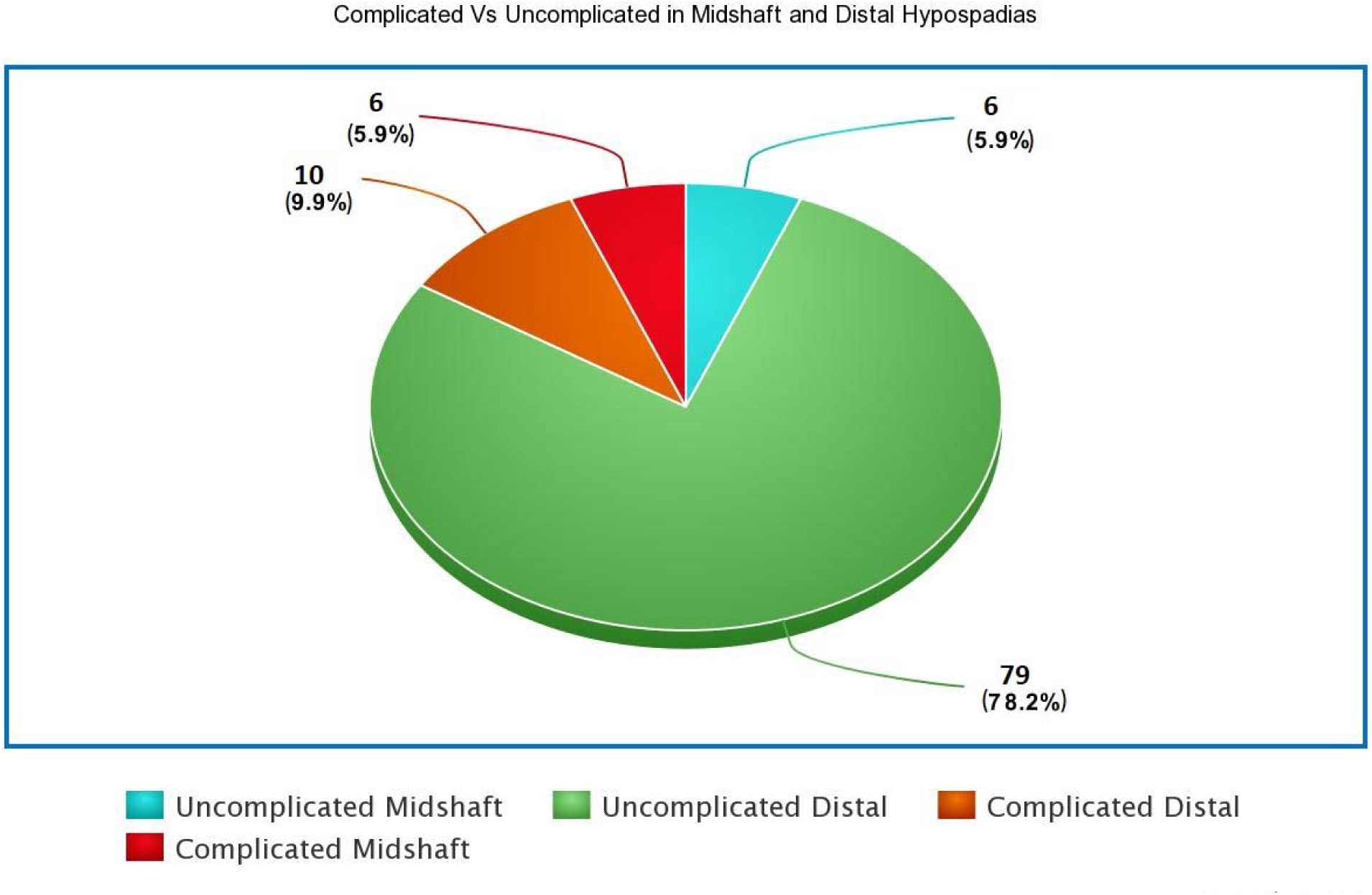
Meta-chart of complicated and uncomplicated percentage in distal and midshaft hypospadias.

**Diagram 5.**
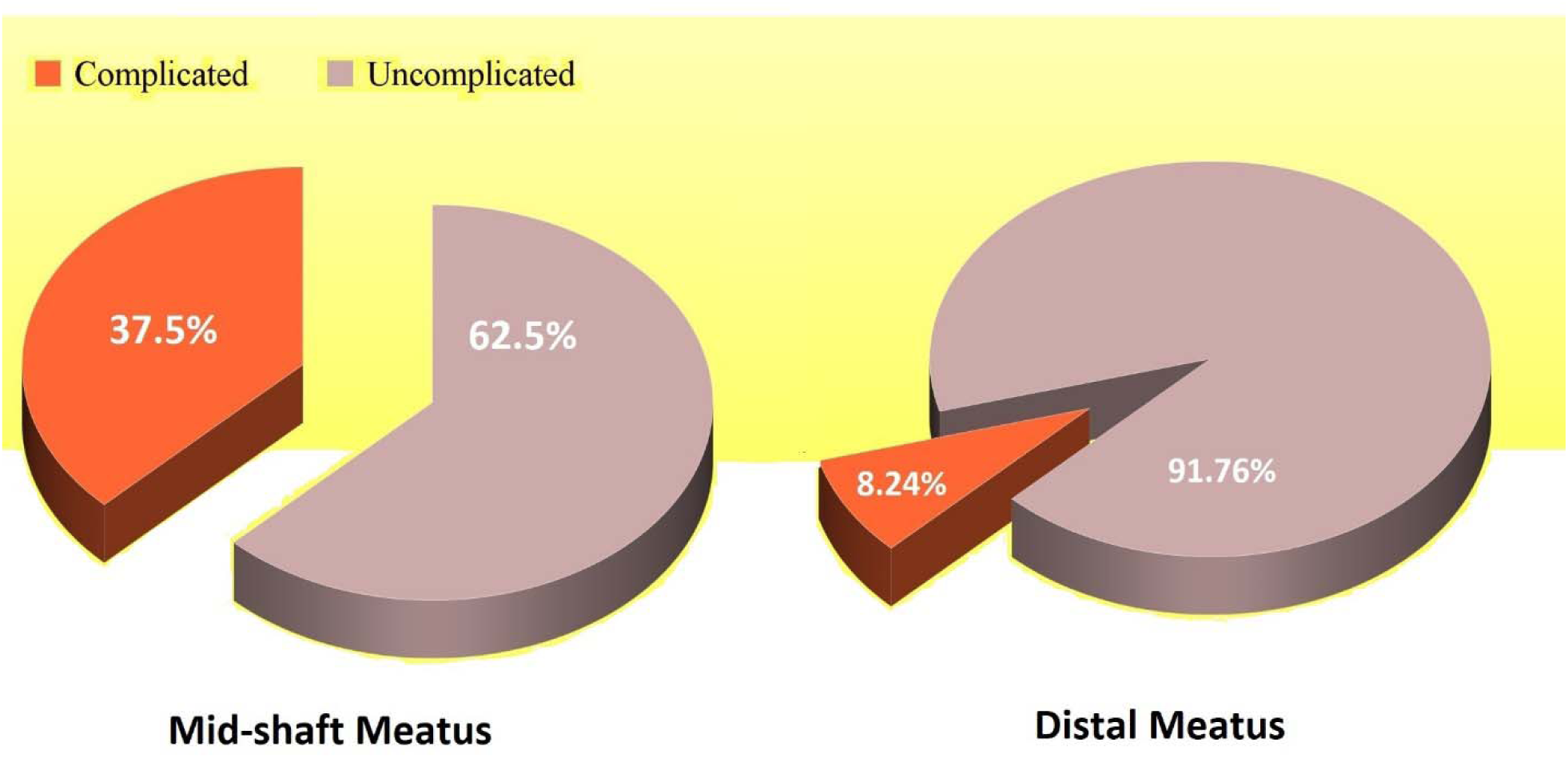
meatal location and complications.

## References

[1] Subramaniam, R., A.F. Spinoit, and P. Hoebeke, Hypospadias repair: an overview of the actual techniques. Seminars in plastic surgery, 2011. 25(3): p. 206–212. https://dx.doi.org/10.1055%2Fs-0031-1281490

[2] Pfistermuller, K.L.M., A.J. McArdle, and P.M. Cuckow, Meta-analysis of complication rates of the tubularized incised plate (TIP) repair. Journal of pediatric urology, 2015. 11(2): p. 54–59. https://doi.org/10.1016/j.jpurol.2014.12.006

[3] Dokter, E.M., et al., Complications after Hypospadias Correction: Prognostic Factors and Impact on Final Clinical Outcome. European journal of pediatric surgery : official journal of Austrian Association of Pediatric Surgery … [et al] = Zeitschrift fur Kinderchirurgie, 2018. 28(2): p. 200–206. https://doi.org/10.1055/s-0037-1599230

[4] Snodgrass, W.T., Re: Effect of suturing technique and urethral plate characteristics on complication rate following hypospadias repair: a prospective randomized study O. Sarhan, M. Saad, T. Helmy and A. Hafez J Urol 2009; 182: 682-686. The Journal of urology, 2010. 183(4): p. 1649–1650. https://doi.org/10.1016/j.juro.2009.12.048

[5] Holland, A.J. and G.H. Smith, Effect of the depth and width of the urethral plate on tubularized incised plate urethroplasty. The Journal of urology, 2000. 164(2): p. 489–491. https://doi.org/10.1016/s0022-5347(05)67408-3

[6] Sarhan, O., et al., Effect of suturing technique and urethral plate characteristics on complication rate following hypospadias repair: a prospective randomized study. The Journal of urology, 2009. 182(2): p. 682–686. https://doi.org/10.1016/j.juro.2009.04.034

[7] Bhat, A. and A.K. Mandal, Acute postoperative complications of hypospadias repair. Indian journal of urology : IJU : journal of the Urological Society of India, 2008. 24(2): p. 241-2.48 https://dx.doi.org/10.4103%2F0970-1591.40622

[8] Mosharafa, A.A., et al., Repair of hypospadias: the effect of urethral plate configuration on the outcome of Duplay-Snodgrass repair. Progres en urologie : journal de l’Association francaise d’urologie et de la Societe francaise d’urologie, 2009. 1 :(7)9p. 507–510. https://doi.org/10.1016/j.purol.2009.02.013

[9] Weber, B.A., et al., Impact of penile degloving and proximal ventral dissection on curvature correction in children with proximal hypospadias. Can Urol Assoc J, 2014. 8(11–12): p. 424–7. https://doi.org/10.5489/cuaj.2337

[10] Bush, N.C., Commentary to ‘Is glans penis width a risk factor for complications after hypospadias repair?’. Journal of pediatric urology, 2016. 12(5): p. 317–318. https://doi.org/10.1016/j.jpurol.2016.05.037

[11] Snodgrass, W.T., C. Granberg, and N.C. Bush, Urethral strictures following urethral plate and proximal urethral elevation during proximal TIP hypospadias repair. Journal of pediatric urology, 2013. 9(6 Pt B): p. 990–994. https://doi.org/10.1016/j.jpurol.2013.04.005

[12] Aslam, R., et al., Medium to long term results following single stage Snodgrass hypospadias repair. Journal of plastic, reconstructive & aesthetic surgery : JPRAS, 2013. 66(11): p. 1591–1595. https://doi.org/10.1016/j.bjps.2013.06.041

[13] Mane, S., J. Arlikar, and N. Dhende, Modified tubularized incised plate urethroplasty. Journal of Indian Association of Pediatric Surgeons, 2013. 18(2): p. 62–65. https://doi.org/10.4103/0971-9261.109354

[14] Xu, N., et al., Comparative outcomes of the tubularized incised plate and transverse island flap onlay techniques for the repair of proximal hypospadias. International urology and nephrology, 2014. 46(3): p. 487–491. http://dx.doi.org/10.1007/s11255-013-0567-z

[15] Springer, A., Assessment of outcome in hypospadias surgery - a review. Frontiers in pediatrics, 2014. 2: p. 2–2. https://dx.doi.org/10.3389%2Ffped.2014.00002

[16] Rynja, S.P., et al., Long-term followup of hypospadias: functional and cosmetic results. The Journal of urology, 2009. 182(4 Suppl): p. 1736–1743. https://doi.org/10.1016/j.juro.2009.03.073

[17] Sarhan, O.M., et al., Factors affecting outcome of tubularized incised plate (TIP) urethroplasty: single-center experience with 500 cases. Journal of pediatric urology, 2009. 5(5): p. 378–382. https://doi.org/10.1016/j.jpurol.2009.02.204

[18] Hansson, E., et al., Analysis of complications after repair of hypospadias. Scandinavian journal of plastic and reconstructive surgery and hand surgery, 2007. 41(3): p. 120–124. https://doi.org/10.1080/02844310701228669

[19] Eassa, W., et al., Risk factors for re-operation following tubularized incised plate urethroplasty: a comprehensive analysis. Urology, 2011. 77(3): p. 716–720. https://doi.org/10.1016/j.urology.2010.07.467

[20] Spinoit, A.-F., et al., Grade of hypospadias is the only factor predicting for re-intervention after primary hypospadias repair: a multivariate analysis from a cohort of 474 patients. Journal of pediatric urology, 2015. 11(2): p. 70.e1-70.e706. https://doi.org/10.1016/j.jpurol.2014.11.014

[21] Arlen, A.M., et al., Further analysis of the Glans-Urethral Meatus-Shaft (GMS) hypospadias score: correlation with postoperative complications. Journal of pediatric urology, 2015. 11(2): p. 71.e1-71.e715. https://doi.org/10.1016/j.jpurol.2014.11.015

[22] Chukwubuike, K.E., et al., Assessment of the effect of urethral plate width on outcome of hypospadias repair. Journal of pediatric urology, 2019. 15(6): p. 627.e1-627.e6. https://doi.org/10.1016/j.jpurol.2019.09.019

[23] Aboutaleb, H., Role of the urethral plate characters in the success of tubularized incised plate urethroplasty. Indian journal of plastic surgery : official publication of the Association of Plastic Surgeons of India, 2014. 47(2): p. 227–231. https://dx.doi.org/10.4103%2F0970-0358.138956

[24] Seleim, H.M., et al., Comprehensive evaluation of grafting the preservable narrow plates with consideration of native plate width at primary hypospadias surgery. Journal of pediatric urology, 2019. 15(4): p. 345.e1-345.e7. https://doi.org/10.1016/j.jpurol.2019.05.002

[25] Nguyen, M.T., W.T. Snodgrass, and M.R. Zaontz, Effect of urethral plate characteristics on tubularized incised plate urethroplasty. The Journal of urology, 2004. 171(3): p. 1260–1262. https://doi.org/10.1097/01.ju.0000110426.32005.91

[26] da Silva, E.A., et al., Role of penile biometric characteristics on surgical outcome of hypospadias repair. Pediatric surgery international, 2014. 30(3): p. 339–344. https://doi.org/10.1016/j.jpurol.2019.09.019

[27] Faasse, M.A., et al., Is glans penis width a risk factor for complications after hypospadias repair? Journal of pediatric urology, 2016. 12(4): p. 202.e1-202.e2025. https://doi.org/10.1016/j.jpurol.2016.04.017

[28] Bush, N.C. and W. Snodgrass, Pre-incision urethral plate width does not impact short-term Tubularized Incised Plate urethroplasty outcomes. Journal of pediatric urology, 2017. 13(6): p. 625.e1.625-e6. https://doi.org/10.1016/j.jpurol.2017.05.020

[29] Qin, D., et al., Application of cavernosum reduction technology in glanuloplasty during repair of moderate-severe hypospadias. Zhongguo xiu fu chong jian wai ke za zhi = Zhongguo xiufu chongjian waike zazhi = Chinese journal of reparative and reconstructive surgery, 2018. 32(11): p. 1454–1457. https://doi.org/10.7507/1002-1892.201801135

[30] El-Karamany, T.M., et al., A Critical Analysis of Stented and Unstented Tubularized Incised Plate Urethroplasty Through a Prospective Randomized Study and Assessment of Factors Influencing the Functional and Cosmetic Outcomes. Urology, 2017. 107: p. 202–208. https://doi.org/10.1016/j.urology.2017.04.056

[31] Bush, N.C., C. Villanueva, and W. Snodgrass, Glans size is an independent risk factor for urethroplasty complications after hypospadias repair. Journal of pediatric urology, 2015. 11(6): p.355.e1-355.e3555. https://doi.org/10.1016/j.jpurol.2015.05.029

[32] Donnahoo, K.K., et al., Etiology, management and surgical complications of congenital chordee without hypospadias. The Journal of urology, 1998. 160(3 Pt 2): p. 1120–1122. https://doi.org/10.1016/S0022-5347(01)62713-7

