## Supplementary figures and images for "A Prospective Study of Outcomes After Tubularized Incised Plate (TIP) Urethroplasty: a Multivariable Analysis of Prognostic Factors in Children"

### Mesurement . suplementary 1

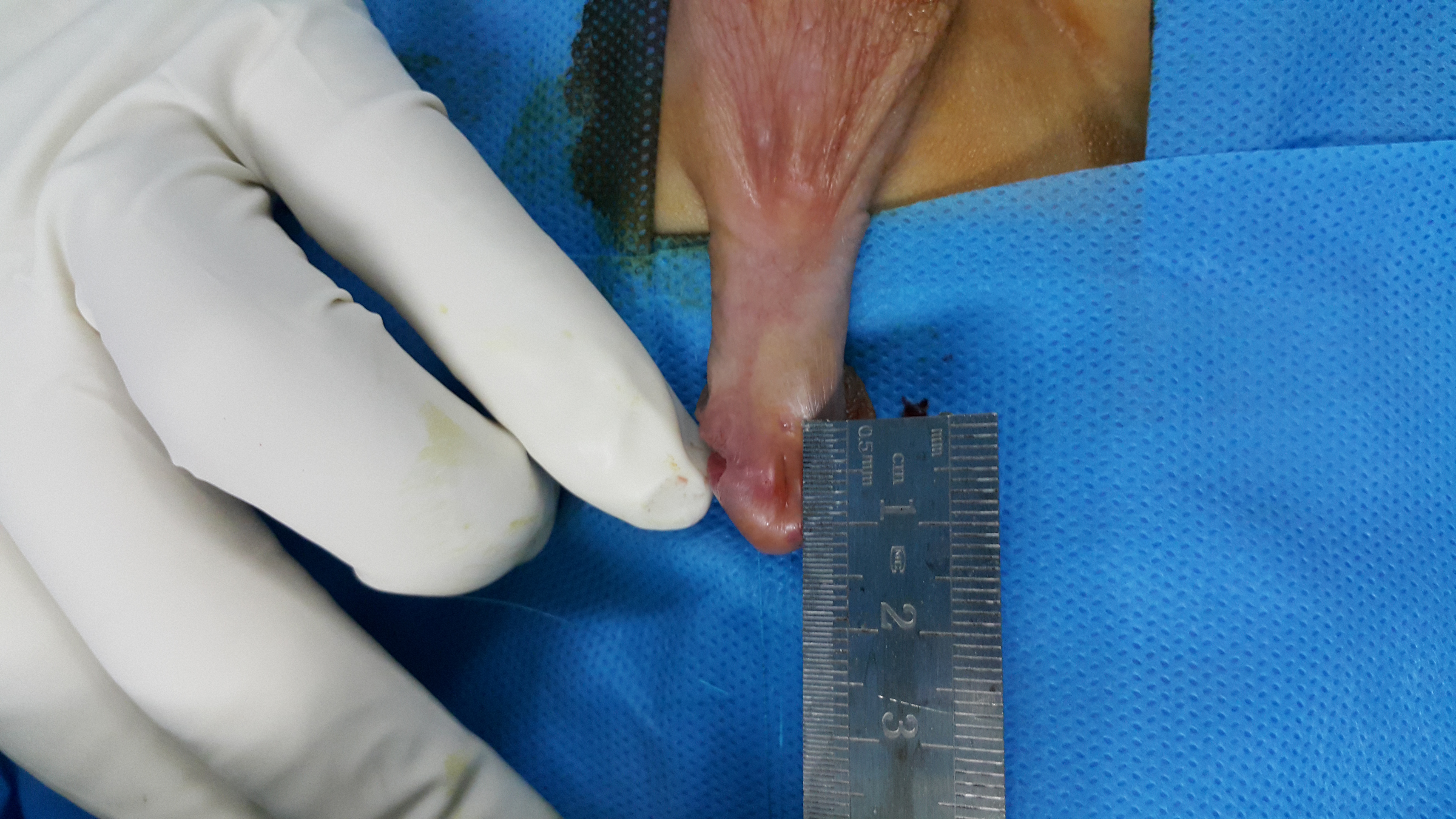

### Mesurement . suplementary 1

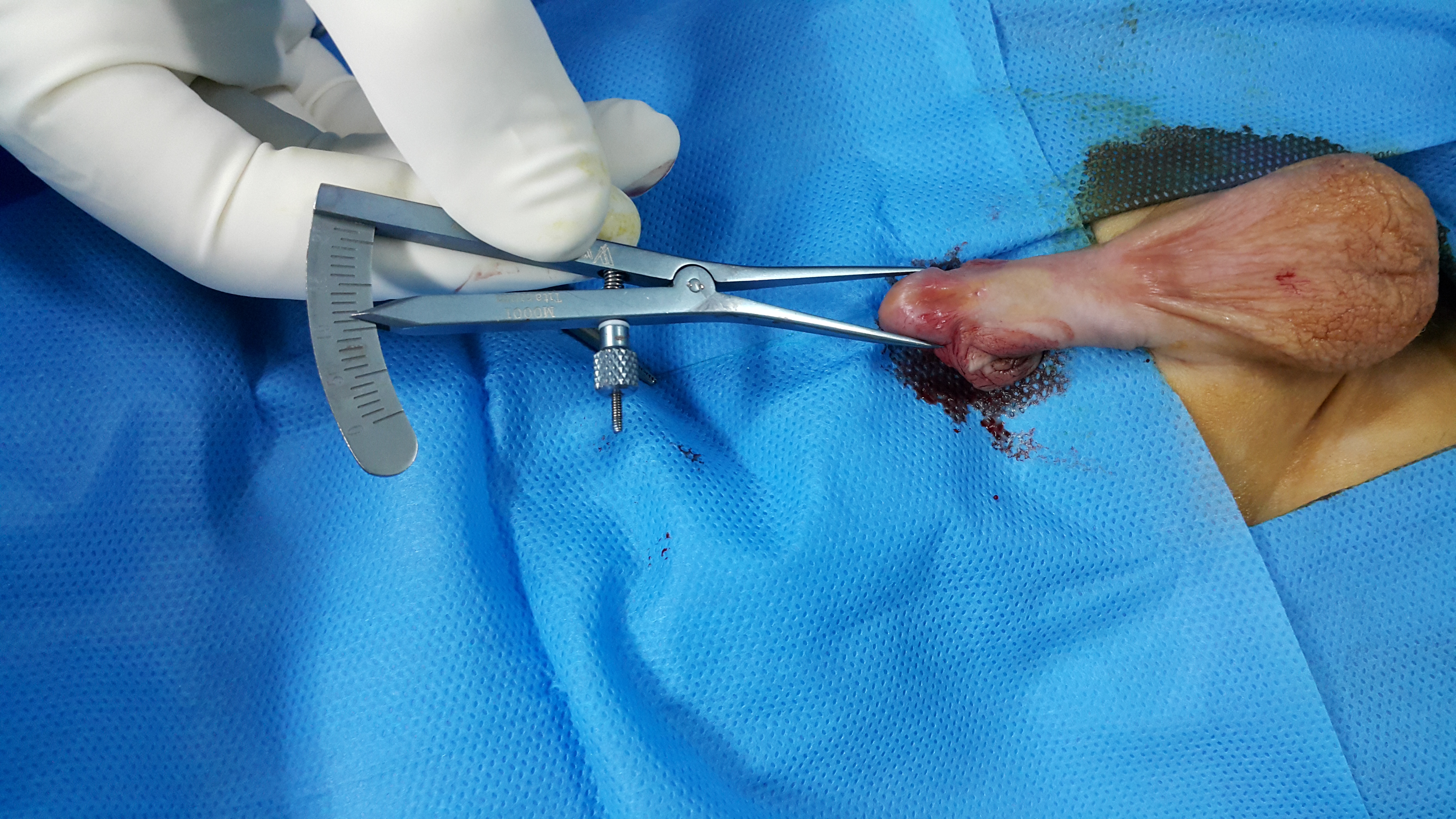

### Mesurement . suplementary 2

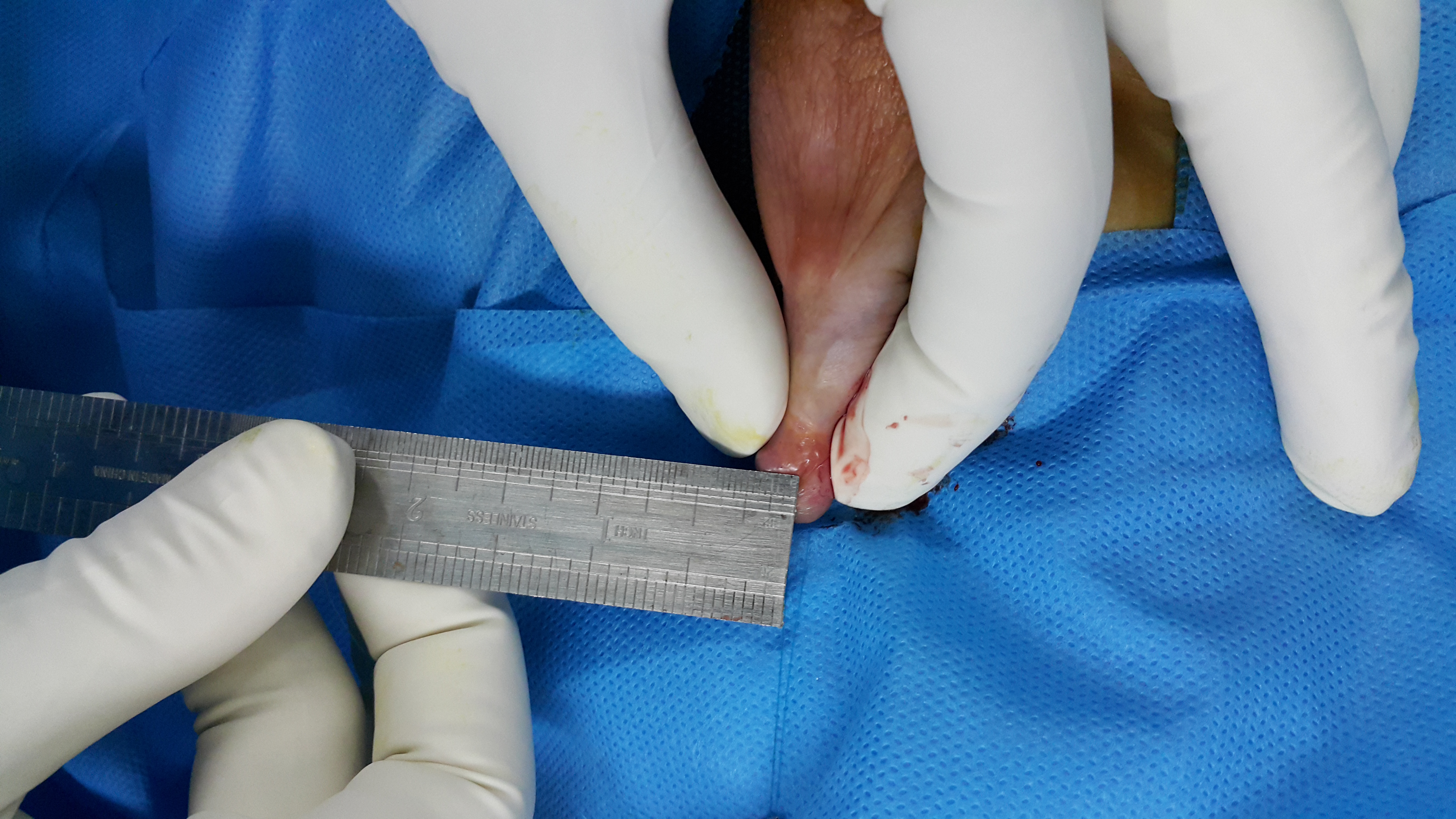
